# Protection of prior natural infection compared to mRNA vaccination against SARS-CoV-2 infection and severe COVID-19 in Qatar

**DOI:** 10.1101/2022.03.17.22272529

**Authors:** Hiam Chemaitelly, Houssein H. Ayoub, Sawsan AlMukdad, Peter Coyle, Patrick Tang, Hadi M. Yassine, Hebah A. Al-Khatib, Maria K. Smatti, Mohammad R. Hasan, Zaina Al-Kanaani, Einas Al-Kuwari, Andrew Jeremijenko, Anvar Hassan Kaleeckal, Ali Nizar Latif, Riyazuddin Mohammad Shaik, Hanan F. Abdul-Rahim, Gheyath K. Nasrallah, Mohamed Ghaith Al-Kuwari, Adeel A. Butt, Hamad Eid Al-Romaihi, Mohamed H. Al-Thani, Abdullatif Al-Khal, Roberto Bertollini, Laith J. Abu-Raddad

## Abstract

**BACKGROUND:** Protection conferred by natural SARS-CoV-2 infection versus COVID-19 vaccination has not been investigated in rigorously controlled studies. We compared head-to-head protection conferred by natural infection to that from the BNT162b2 (Pfizer-BioNTech) and mRNA-1273 (Moderna) vaccines in Qatar, between February 28, 2020 and March 6, 2022.

**METHODS:** Two national matched retrospective target-trial cohort studies were conducted to compare incidence of SARS-CoV-2 infection and COVID-19 hospitalization and death among those with a documented primary infection to incidence among those with a two-dose primary-series vaccination. Associations were estimated using Cox proportional-hazards regression models.

**RESULTS:** The overall adjusted hazard ratio (AHR) for infection was 0.46 (95% CI: 0.45-0.48) comparing those with a prior infection to those vaccinated with BNT162b2, and 0.51 (95% CI: 0.48-0.53) comparing those with a prior infection to those vaccinated with mRNA-1273. For BNT162b2, the AHR decreased gradually from 0.55 (95% CI: 0.46-0.65) in the fourth month after primary infection/vaccination to 0.31 (95% CI: 0.27-0.37) in the eighth month, while for mRNA-1273, it decreased from 0.80 (95% CI: 0.59-1.07) to 0.35 (95% CI: 0.29-0.41) over the same time period. During the Omicron wave, the AHR was ∼0.50 for BNT162b2 and ∼0.60 for mRNA-1273. The overall AHR for any severe, critical, or fatal COVID-19 (against all variants) was 0.32 (95% CI: 0.10-1.00) for BNT162b2, and 0.58 (95% CI: 0.14-2.43) for mRNA-1273.

**CONCLUSIONS:** Natural infection was associated with stronger and more durable protection against infection, regardless of the variant, than mRNA primary-series vaccination. Nonetheless, vaccination remains the safest and optimal tool of protection against infection and COVID-19 hospitalization and death.

## Introduction

Coronavirus disease 2019 (COVID-19) vaccines induce protection against severe acute respiratory syndrome coronavirus 2 (SARS-CoV-2) infection and COVID-19 hospitalization and death.^1-4^ Natural infection with SARS-CoV-2 also induces protection against subsequent SARS-CoV-2 infection and COVID-19 hospitalization and death.^5,6^ A growing number of studies suggests that there are differences in the level and durability of protection of natural infection versus vaccination.^7-11^ These differences may arise from differences in the mechanism of action,^12,13^ from mucosal immunity,^14,15^ volume and nature of neutralizing antibody titers,^12,16,17^ or circulating variants.^18-22^

Protection conferred by natural infection versus vaccination has not been investigated in rigorously controlled studies that directly compare them. In this study, we compared protection conferred by natural infection to that conferred by the BNT162b2^1^ (Pfizer-BioNTech) and mRNA-1273^2^ (Moderna) vaccines in Qatar, using a matched target-trial cohort study design.^23,24^ The study is basically a retrospective, unblinded “controlled trial”^23,24^ that allows a head-to-head comparison of protection provoked by natural infection and vaccination against SARS-CoV-2 infection and against COVID-19 hospitalization and death.

## Methods

### Study population and data sources

This study was conducted in the resident population of Qatar. It analyzed the national, federated databases for COVID-19 vaccination, laboratory testing, hospitalization, and death, retrieved from the integrated nationwide digital-health information platform. Databases include all SARS-CoV-2-related data and associated demographic information, with no missing information, since pandemic onset—such as all polymerase chain reaction (PCR) tests and more recently, rapid antigen tests conducted at healthcare facilities (from January 5, 2022 onward).

Every PCR test (but not rapid antigen tests) conducted in Qatar is classified on the basis of symptoms and the reason for testing (clinical symptoms, contact tracing, surveys or random testing campaigns, individual requests, routine healthcare testing, pre-travel, at port of entry, or other). Qatar has unusually young, diverse demographics, in that only 9% of its residents are ≥50 years of age, and 89% are expatriates from over 150 countries.^25^ Qatar launched its COVID-19 vaccination program at the end of December of 2020 using both the BNT162b2 and mRNA-1273 mRNA vaccines.^26^ Nearly all individuals were vaccinated in Qatar, but if vaccinated elsewhere, those vaccinations were still recorded in the health system at the port of entry upon arrival in Qatar. Further descriptions of the study population and these national databases have been reported previously.^3,18-22,25^

### Study design

Durability of protection from natural infection was compared to that of primary series of two-dose vaccination, for both the BNT162b2 and mRNA-1273 vaccines, using two matched, retrospective cohort studies that emulated a target trial.^23,24^ Incidence of reinfection in the cohort of individuals with a documented SARS-CoV-2 primary (first) infection (natural-infection cohort) was compared to incidence of infection in the cohort of vaccinated individuals who have not yet received their third (booster) dose (vaccinated cohort). Documentation of infection was based on positive PCR or rapid-antigen tests. Laboratory methods for real-time, reverse-transcription PCR (RT-qPCR) testing and rapid antigen testing are found in Supplementary Appendix Section S1.

Any individual with a documented primary infection from pandemic onset in Qatar on February 28, 2020 up to March 6, 2022 (end of study) was eligible for inclusion in the natural-infection cohort, provided that the individual had no vaccination record before the start of follow-up. Any individual with at least two vaccine doses between January 5, 2021 (date of first second-dose vaccination in Qatar) and March 6, 2022 was eligible for inclusion in either the BNT162b2 or mRNA-1273 vaccinated cohorts, provided that the individual had no record of a prior documented infection and had not received a booster vaccination before the start of follow-up.

Individuals in the natural-infection cohort were exact-matched in a one-to-one ratio by sex, 10-year age group, and nationality, to individuals in each of the vaccinated cohorts, to control for known differences in the risk of exposure to SARS-CoV-2 infection in Qatar.^25,27-30^ Matching by these factors was shown previously to provide adequate control of differences in the risk of exposure to the infection in Qatar in studies of different epidemiologic designs and that included control groups, including target-trial cohort studies.^3,4,19,26,31^

Matching was performed through an iterative process to ensure that each individual in the vaccinated cohorts was alive, infection-free, and had not received a booster dose at the start of follow-up. Onset of symptoms for SARS-CoV-2 infection occurs several days after acquisition of the virus.^32^ Accordingly, to control for time since inducement of SARS-CoV-2 immunity, and to control for epidemic phase and variant exposure throughout time of follow-up, each individual in the vaccinated cohorts was matched to an individual in the natural-infection cohort who had a documented primary infection within a week after the vaccinated match received the first dose.

SARS-CoV-2 reinfection is conventionally defined as a documented infection ≥90 days after an earlier infection, to avoid misclassification of prolonged PCR positivity as reinfection, if a shorter time interval is used.^6,8,9^ Therefore, each matched pair was followed from the day the individual in the natural-infection cohort completed 90 days since the documented primary infection.

Both members of each matched pair were censored on the date that the individual in the natural-infection cohort received the first vaccine dose, or the individual in the vaccinated cohort received a booster dose, to ensure exchangeability.^24,33^ Accordingly, individuals were followed up until the first of any of the following events: a documented SARS-CoV-2 infection (defined as the first PCR-positive or rapid-antigen-positive test after the start of follow-up, regardless of symptoms or reason for testing), or first-dose vaccination of the individual in the natural-infection cohort (with matched pair censoring), or booster-dose vaccination of the individual in the vaccinated cohort (with matched pair censoring), or death, or end of study censoring (March 6, 2022).

### Study outcomes

The primary outcome of the study was occurrence of a documented SARS-CoV-2 infection during follow-up, regardless of symptoms or reason for testing. Moreover, we investigated, as a secondary outcome, occurrence of any severe,^34^ critical,^34^ or fatal^35^ COVID-19. Classification of COVID-19 case severity (acute-care hospitalizations),^34^ criticality (intensive-care-unit hospitalizations),^34^ and fatality^35^ followed World Health Organization guidelines. Assessments were made by trained medical personnel independent of study investigators and using individual chart reviews, as part of a national protocol applied to every hospitalized COVID-19 patient. Details of COVID-19 severity, criticality, and fatality classifications are found in Section S2.

Every hospitalized COVID-19 patient underwent an infection severity assessment every three days until discharge or death. We classified individuals who progressed to severe, critical, or fatal COVID-19 between the time of the documented infection and the end of the study based on their worst outcome, starting with death,^35^ followed by critical disease,^34^ and then severe disease.^34^

### Statistical analysis

Eligible and matched cohorts were described using frequency distributions and measures of central tendency, and compared using standardized mean differences (SMDs). An SMD of <0.1 indicated adequate matching.^36^ Cumulative incidence of infection (defined as the proportion of individuals at risk, whose primary endpoint during follow-up was a reinfection for the natural-infection cohort, or an infection for the vaccinated cohort) was estimated in each cohort using the Kaplan–Meier estimator method.^37^ Incidence rate of infection in each cohort, defined as the number of identified infections divided by the number of person-weeks contributed by all individuals in the cohort, was estimated, along with its 95% confidence interval (CI), using a Poisson log-likelihood regression model with the STATA 17.0 *stptime* command.

The hazard ratio, comparing incidence of infection in both cohorts and the corresponding 95% CIs, was calculated using Cox regression adjusted for the matching factors with the STATA 17.0 *stcox* command. Schoenfeld residuals and log-log plots for survival curves were used to test the proportional-hazards assumption and to investigate its adequacy. 95% CIs were not adjusted for multiplicity; thus, they should not be used to infer definitive differences between cohorts.

Interactions were not considered. Subgroup analyses were conducted to estimate adjusted hazard ratio stratified by month of follow-up. Statistical analyses were conducted using STATA/SE version 17.0 (Stata Corporation, College Station, TX, USA).

### Oversight

Hamad Medical Corporation and Weill Cornell Medicine-Qatar Institutional Review Boards approved this retrospective study with waiver of informed consent. The study was reported following the Strengthening the Reporting of Observational studies in Epidemiology (STROBE) guidelines. The STROBE checklist is found in Table S1.

## Results

### Study population

Between February 28, 2020 and March 6, 2022, 796,938 individuals had a PCR-confirmed or rapid-antigen-confirmed primary infection, of whom 515,705 individuals were unvaccinated at the time of diagnosis. The median date of the infection was March 20, 2021.

Between January 5, 2021 and March 6, 2022, 1,313,588 individuals received at least two BNT162b2 doses, and 367,266 of these received a booster dose. The median date was May 4, 2021 for the first dose, May 24, 2021 for the second dose, and December 29, 2021 for the booster dose. The median time elapsed between the first and second doses was 21 days (interquartile range (IQR), 21-22 days), and between the second and booster doses was 252 days (IQR, 233-276 days).

Between January 24, 2021 and March 6, 2022, 894,553 individuals received at least two mRNA-1273 doses, and 156,795 of these received a booster dose. The median date was May 28, 2021 for the first dose, June 27, 2021 for the second dose, and January 18, 2022 for the booster dose. The median time elapsed between the first and second doses was 28 days (IQR, 28-30 days), and between the second and booster doses was 236 days (IQR, 212-262 days).

The process that was used to select the study populations of the two cohort studies is shown in Figure 1. Characteristics of the study populations are described in Table 1. Figure S1 shows the distribution of documented primary infections and of first-dose vaccinations by calendar month in the matched cohorts. While first-dose BNT162b2 vaccinations tended to be more broadly distributed over time, most first-dose mRNA-1273 vaccinations occurred in March-April of 2021. Figure S2 shows the distribution of the durations of follow-up in these cohorts.

**Figure 1.**
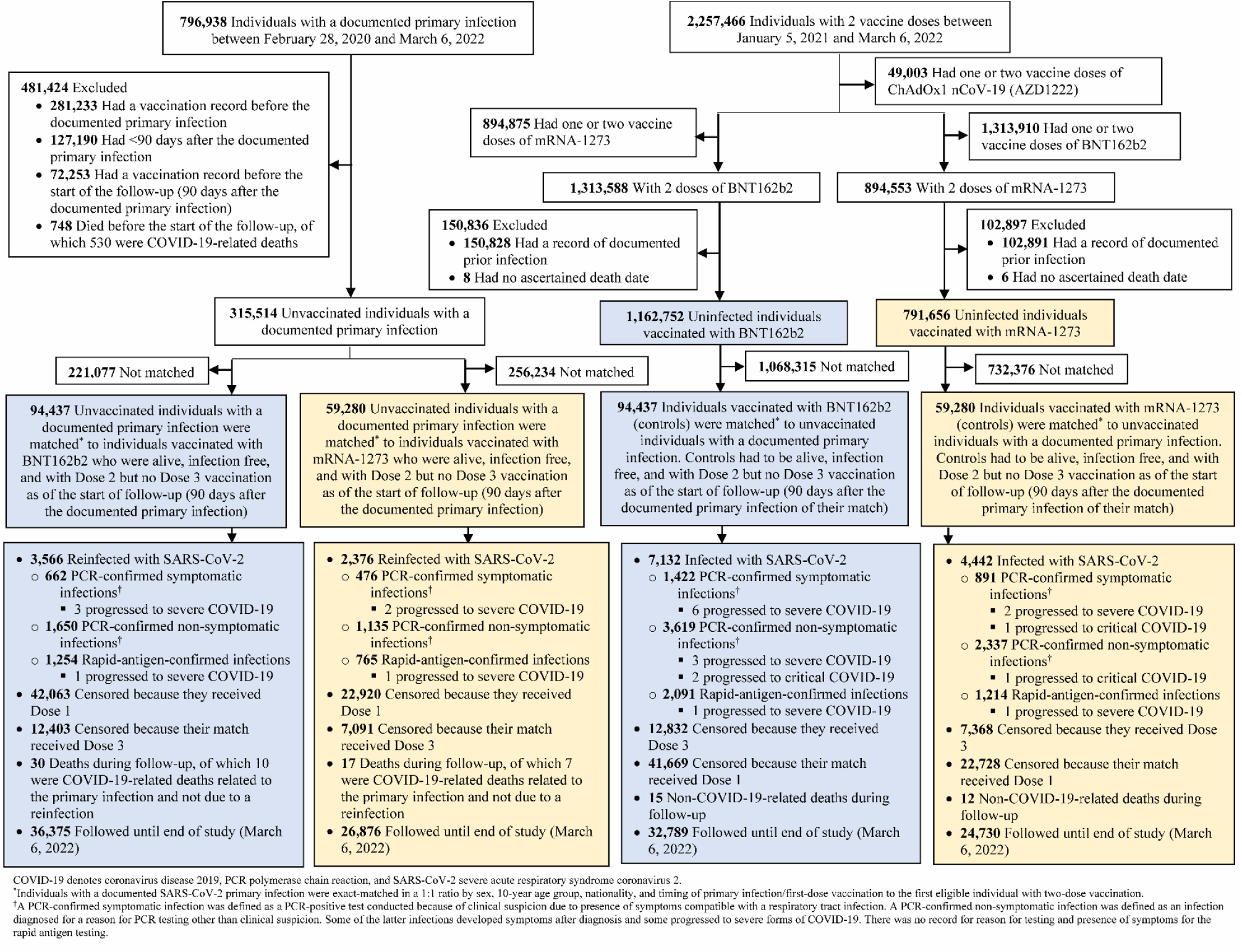
Cohort selection for investigating protection resulting from natural infection compared to that of BNT162b2 and mRNA-1273 vaccination, against incidence of SARS-CoV-2 infection and COVID-19 hospitalization and death.

**Table 1.** Baseline characteristics of the eligible and matched natural-infection cohort and the BNT162b2- and mRNA-1273-vaccinated cohorts.

| Characteristics | Natural infection versus BNT162b2 vaccination study |  |  |  |  |  | Natural infection versus mRNA-1273 vaccination study |  |  |  |  |  |
| --- | --- | --- | --- | --- | --- | --- | --- | --- | --- | --- | --- | --- |
|  | Full eligible cohorts |  |  | Matched cohorts<sup>†</sup> |  |  | Full eligible cohorts |  |  | Matched cohorts<sup>‡</sup> |  |  |
|  | Natural infection | BNT162b2 primary-series vaccination | SMD<sup>†</sup> | Natural infection | BNT162b2 primary-series vaccination | SMD<sup>†</sup> | Natural infection | mRNA-1273 primary-series vaccination | SMD<sup>†</sup> | Natural infection | mRNA-1273 primary-series vaccination | SMD<sup>†</sup> |
|  | N=315,514 | N=1,162,752 |  | N=94,437 | N=94,437 |  | N=315,514 | N=791,656 |  | N=59,280 | N=59,280 |  |
| Median age (IQR) — years | 32 (24-40) | 36 (28-44) | 0.40<sup>‡</sup> | 33 (26-40) | 33 (27-40) | 0.06<sup>‡</sup> | 32 (24-40) | 35 (30-42) | 0.44<sup>‡</sup> | 33 (28-40) | 33 (28-40) | 0.05<sup>‡</sup> |
| Age group — no. (%) |  |  |  |  |  |  |  |  |  |  |  |  |
| 0-9 years | 33,134 (10.5) | 1,766 (0.2) |  | 1 (<0.01) | 1 (<0.01) |  | 33,134 (10.5) | 0 (0.0) |  | -- | -- |  |
| 10-19 years | 24,987 (7.9) | 116,858 (10.1) |  | 9,874 (10.5) | 9,874 (10.5) |  | 24,987 (7.9) | 5,196 (0.7) |  | 2,631 (4.4) | 2,631 (4.4) |  |
| 20-29 years | 74,821 (23.7) | 213,181 (18.3) |  | 24,875 (26.3) | 24,875 (26.3) |  | 74,821 (23.7) | 184,908 (23.4) |  | 17,905 (30.2) | 17,905 (30.2) |  |
| 30-39 years | 99,674 (31.6) | 397,062 (34.2) | 0.53 | 33,834 (35.8) | 33,834 (35.8) | 0.00 | 99,674 (31.6) | 338,873 (42.8) | 0.66 | 22,815 (38.5) | 22,815 (38.5) | 0.00 |
| 40-49 years | 54,085 (17.1) | 256,805 (22.1) |  | 18,032 (19.1) | 18,032 (19.1) |  | 54,085 (17.1) | 185,907 (23.5) |  | 11,442 (19.3) | 11,442 (19.3) |  |
| 50-59 years | 21,106 (6.7) | 118,447 (10.2) |  | 5,916 (6.3) | 5,916 (6.3) |  | 21,106 (6.7) | 62,572 (7.9) |  | 3,444 (5.8) | 3,444 (5.8) |  |
| 60-69 years | 6,044 (1.9) | 44,526 (3.8) |  | 1,464 (1.6) | 1,464 (1.6) |  | 6,044 (1.9) | 11,997 (1.5) |  | 801 (1.4) | 801 (1.4) |  |
| 70+ years | 1,663 (0.5) | 14,107 (1.2) |  | 441 (0.5) | 441 (0.5) |  | 1,663 (0.5) | 2,203 (0.3) |  | 242 (0.4) | 242 (0.4) |  |
| Sex |  |  |  |  |  |  |  |  |  |  |  |  |
| Male | 229,090 (72.6) | 804,317 (69.2) | 0.08 | 64,282 (68.1) | 64,282 (68.1) | 0.00 | 229,090 (72.6) | 640,327 (80.9) | 0.20 | 40,945 (69.1) | 40,945 (69.1) | 0.00 |
| Female | 86,424 (27.4) | 358,435 (30.8) |  | 30,155 (31.9) | 30,155 (31.9) |  | 86,424 (27.4) | 151,329 (19.1) |  | 18,335 (30.9) | 18,335 (30.9) |  |
| Nationality<sup>§</sup> |  |  |  |  |  |  |  |  |  |  |  |  |
| Bangladeshi | 27,415 (8.7) | 128,393 (11.0) |  | 6,297 (6.7) | 6,297 (6.7) |  | 27,415 (8.7) | 153,424 (19.4) |  | 4,009 (6.8) | 4,009 (6.8) |  |
| Egyptian | 16,578 (5.3) | 65,273 (5.6) |  | 4,196 (4.4) | 4,196 (4.4) |  | 16,578 (5.3) | 31,117 (3.9) |  | 1,987 (3.4) | 1,987 (3.4) |  |
| Filipino | 25,125 (8.0) | 107,574 (9.3) |  | 11,557 (12.2) | 11,557 (12.2) |  | 25,125 (8.0) | 70,650 (8.9) |  | 7,829 (13.2) | 7,829 (13.2) |  |
| Indian | 82,954 (26.3) | 259,031 (22.3) |  | 29,237 (31.0) | 29,237 (31.0) |  | 82,954 (26.3) | 218,399 (27.6) |  | 20,261 (34.2) | 20,261 (34.2) |  |
| Nepalese | 37,512 (11.9) | 96,910 (8.3) | 0.20 | 7,977 (8.5) | 7,977 (8.5) | 0.00 | 37,512 (11.9) | 109,829 (13.9) | 0.52 | 5,265 (8.9) | 5,265 (8.9) | 0.00 |
| Pakistani | 17,487 (5.5) | 49,843 (4.3) |  | 5,034 (5.3) | 5,034 (5.3) |  | 17,487 (5.5) | 42,569 (5.4) |  | 3,405 (5.7) | 3,405 (5.7) |  |
| Qatari | 38,611 (12.2) | 160,941 (13.8) |  | 9,873 (10.5) | 9,873 (10.5) |  | 38,611 (12.2) | 14,594 (1.8) |  | 4,533 (7.7) | 4,533 (7.7) |  |
| Sri Lankan | 10,029 (3.2) | 35,340 (3.0) |  | 2,916 (3.1) | 2,916 (3.1) |  | 10,029 (3.2) | 32,504 (4.1) |  | 1,933 (3.3) | 1,933 (3.3) |  |
| Sudanese | 8,586 (2.7) | 25,820 (2.2) |  | 2,420 (2.6) | 2,420 (2.6) |  | 8,586 (2.7) | 13,540 (1.7) |  | 1,232 (2.1) | 1,232 (2.1) |  |
| Other nationalities<sup>¶</sup> | 51,217 (16.2) | 233,627 (20.1) |  | 14,930 (15.8) | 14,930 (15.8) |  | 51,217 (16.2) | 105,030 (13.3) |  | 8,826 (14.9) | 8,826 (14.9) |  |

IQR denotes interquartile range and SMD standardized mean difference.
<sup>†</sup>Individuals with a documented SARS-CoV-2 primary infection were exact-matched in a 1:1 ratio by sex, 10-year age group, nationality, and timing of primary infection/first-dose vaccination to the first eligible individual with two-dose vaccination.
<sup>‡</sup>SMD is the difference in the mean of a covariate between groups divided by the pooled standard deviation. An SMD <0.1 indicates adequate matching.
<sup>§</sup>SMD is for the mean difference between groups divided by the pooled standard deviation.
<sup>¶</sup>Nationalities were chosen to represent the most populous groups in Qatar.
<sup>¶</sup>These comprise 172 other nationalities in individuals with natural infection and 190 other nationalities in individuals with BNT162b2 vaccination in the unmatched cohorts of natural infection versus BNT162b2 vaccination study, and 116 other nationalities in individuals with natural infection and 116 other nationalities in individuals with BNT162b2 vaccination in the matched cohorts of natural infection versus BNT162b2 vaccination study. These also comprise 172 other nationalities in individuals with natural infection and 173 other nationalities in individuals with mRNA-1273 vaccination in the unmatched cohorts of natural infection versus mRNA-1273 vaccination study, and 102 other nationalities in individuals with natural infection and 102 other nationalities in individuals with mRNA-1273 vaccination in the matched cohorts of natural infection versus mRNA-1273 vaccination study.

The two cohort studies were based on the total population of Qatar; thus, study populations are broadly representative of the internationally diverse, but young and predominantly male, total population of Qatar (Table S2).

### Natural infection versus BNT162b2 vaccination

The median time of follow-up was 108 days (IQR, 32-202 days) for the natural-infection cohort and 107 days (IQR, 32-197 days) for the BNT162b2-vaccinated cohort (Figure 2A). A total of 3,566 reinfections were recorded in the natural-infection cohort during follow-up (Figure 1). Four of these reinfections progressed to severe COVID-19, but none to critical or fatal COVID-19. A total of 7,132 infections were recorded in the BNT162b2-vaccinated cohort. Of these, 10 progressed to severe, 2 to critical, but none to fatal COVID-19.

**Figure 2.**
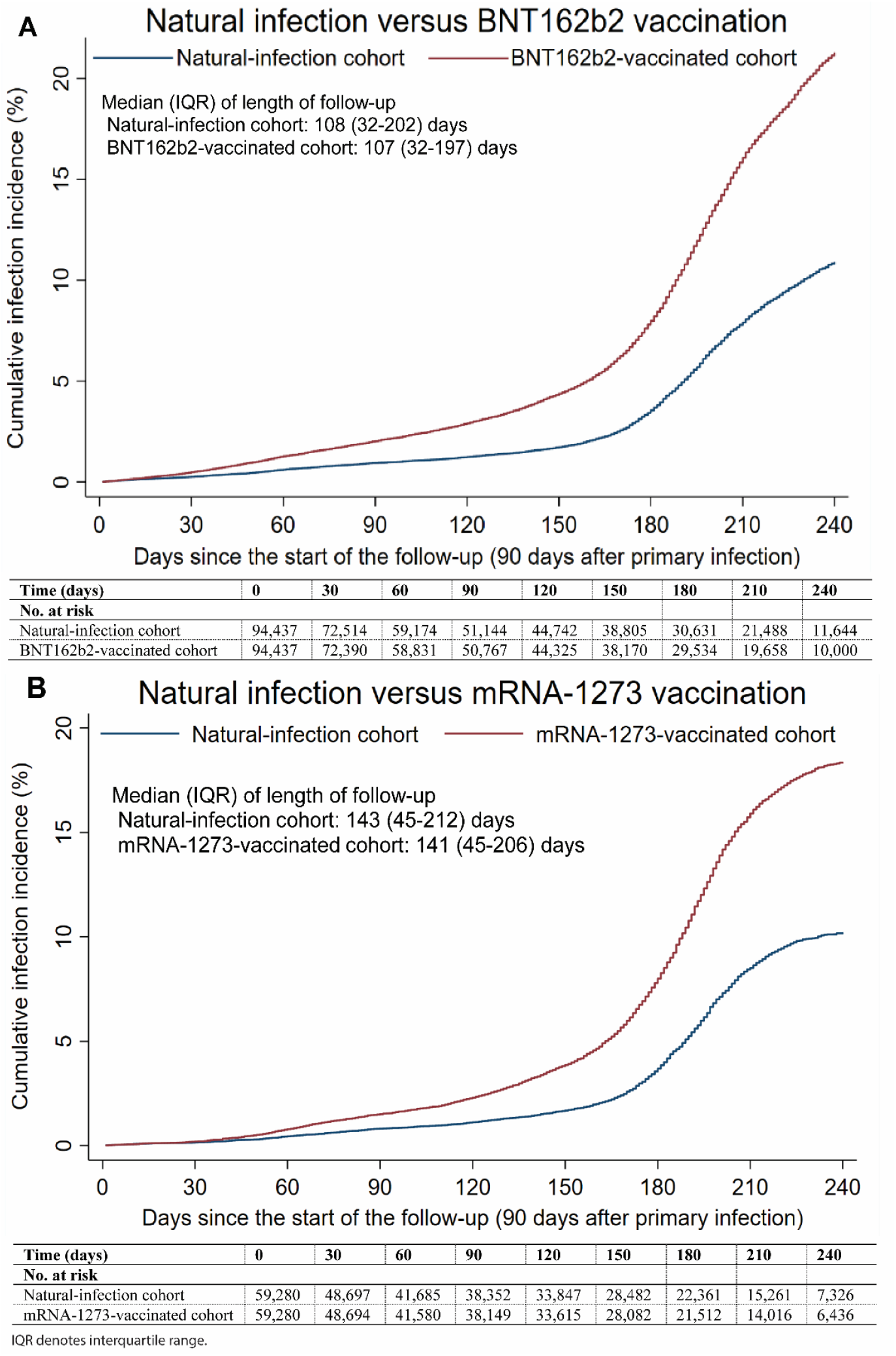
Cumulative incidence of infection in the natural-infection cohort and in the (A) BNT162b2-vaccinated cohort and (B) the mRNA-1273-vaccinated cohort.

Cumulative incidence of infection was estimated at 10.9% (95% CI: 10.5-11.3%) for the natural-infection cohort and at 21.3% (95% CI: 20.8-21.8%) for the BNT162b2-vaccinated cohort, 240 days after the start of follow-up (Figure 2A). Incidence was limited in the natural-infection cohort until day 160 of follow-up, but then increased rapidly with the onset of the Omicron (B.1.1.529)^38^ variant wave on December 19, 2021, which peaked in mid-January of 2022.^6,24,39,40^ Prior to day 160, incidence was dominated by the Alpha (B.1.1.7),^38^ Beta (B.1.351),^38^ and Delta (B.1.617.2)^38^ variants.^3,20,39,41,42^ Meanwhile, there was considerable incidence in the BNT162b2-vaccinated cohort prior to the Omicron wave, and much larger incidence after the onset of this wave.

The overall hazard ratio for infection, adjusted for sex, 10-year age group, 10 nationality groups, and time since primary infection/vaccination, was estimated at 0.46 (95% CI: 0.45-0.48; Table 2). However, the adjusted hazard ratio varied by month of follow-up (Table 3). It was 0.55 (95% CI: 0.46-0.65) in the first month of follow-up, that is in the fourth month after primary infection/vaccination, but decreased gradually to 0.31 (95% CI: 0.27-0.37) in the eighth month after primary infection/vaccination. Nonetheless, the adjusted hazard ratio increased after the onset of the Omicron wave. It was ∼0.50 during the time of follow-up that coincided with this wave.

**Table 2.** Hazard ratios for the incidence of SARS-CoV-2 infection and incidence of COVID-19 hospitalization and death, comparing the natural-infection cohort to the BNT162b2- and mRNA-1273-vaccinated cohorts.

| Epidemiological measure | Natural infection versus BNT162b2 vaccination study |  | Natural infection versus mRNA-1273 vaccination study |  |
| --- | --- | --- | --- | --- |
|  | Natural-infection cohort | BNT162b2-vaccinated cohort | Natural-infection cohort | mRNA-1273-vaccinated cohort |
| Total follow-up time (person-weeks) | 1,619,572 | 1,588,884 | 1,131,718 | 1,114,485 |
| Incidence rate of infection (per 10,000 person-weeks) | 22.0 (21.3 to 22.8) | 44.9 (43.9 to 45.9) | 21.0 (20.2 to 21.9) | 39.9 (38.7 to 41.1) |
| Unadjusted hazard ratio for SARS-CoV-2 infection (95% CI) | 0.47 (0.46 to 0.49) |  | 0.51 (0.49 to 0.54) |  |
| Adjusted hazard ratio for SARS-CoV-2 infection* (95% CI) | 0.46 (0.45 to 0.48) |  | 0.51 (0.48 to 0.53) |  |
| Unadjusted hazard ratio for severe, critical, or fatal COVID-19† (95% CI) | 0.32 (0.10 to 0.99) |  | 0.58 (0.14 to 2.41) |  |
| Adjusted hazard ratio for severe, critical, or fatal COVID-19*† (95% CI) | 0.32 (0.10 to 1.00) |  | 0.58 (0.14 to 2.43) |  |

CI denotes confidence interval, COVID-19 coronavirus disease 2019, and SARS-CoV-2 severe acute respiratory syndrome coronavirus 2.
\*Cox regression analysis adjusted for sex, 10-year age group (Table 1), 10 nationality groups (Table 1), and timing of primary infection/first dose vaccination.
†Severity,<sup>34</sup> criticality,<sup>34</sup> and fatality<sup>35</sup> were defined according to the World Health Organization guidelines.

**Table 3.** **Hazard ratios for incidence of SARS-CoV-2 infection month-by-month since the start of follow-up (90 days after primary infection/first-dose vaccination) comparing the natural-infection cohort to the BNT162b2- and mRNA-1273-vaccinated cohorts.**

| Months* | Natural-infection cohort† |  | mRNA-vaccinated cohort† |  | Unadjusted hazard ratio<br>(95% CI) | Adjusted hazard ratio<br>(95% CI)‡ |
| --- | --- | --- | --- | --- | --- | --- |
|  | N | Cumulative incidence<br>(95% CI) | N | Cumulative incidence<br>(95% CI) |  |  |
| Natural infection versus BNT162b2 vaccination study§ |  |  |  |  |  |  |
| Month 1 of follow-up (Month 4 after primary infection/first-dose vaccination) | 72,514 | 0.26 (0.23 to 0.30) | 72,390 | 0.49 (0.45 to 0.54) | 0.55 (0.47 to 0.66) | 0.55 (0.46 to 0.65) |
| Month 2 of follow-up (Month 5 after primary infection/first-dose vaccination) | 59,174 | 0.61 (0.56 to 0.67) | 58,831 | 1.28 (1.20 to 1.36) | 0.44 (0.38 to 0.52) | 0.43 (0.37 to 0.50) |
| Month 3 of follow-up (Month 6 after primary infection/first-dose vaccination) | 51,144 | 0.95 (0.87 to 1.02) | 50,767 | 2.02 (1.92 to 2.14) | 0.45 (0.38 to 0.53) | 0.43 (0.36 to 0.51) |
| Month 4 of follow-up (Month 7 after primary infection/first-dose vaccination) | 44,742 | 1.24 (1.16 to 1.33) | 44,325 | 2.91 (2.77 to 3.05) | 0.32 (0.27 to 0.39) | 0.31 (0.26 to 0.38) |
| Month 5 of follow-up (Month 8 after primary infection/first-dose vaccination) | 38,805 | 1.72 (1.62 to 1.84) | 38,170 | 4.38 (4.20 to 4.56) | 0.33 (0.28 to 0.38) | 0.31 (0.27 to 0.37) |
| Month 6 of follow-up (Month 9 after primary infection/first-dose vaccination) | 30,631 | 3.56 (3.38 to 3.74) | 29,534 | 7.99 (7.73 to 8.25) | 0.47 (0.43 to 0.52) | 0.46 (0.42 to 0.51) |
| Month 7 of follow-up (Month 10 after primary infection/first-dose vaccination) | 21,488 | 7.91 (7.62 to 8.21) | 19,658 | 16.06 (15.66 to 16.46) | 0.51 (0.48 to 0.55) | 0.50 (0.47 to 0.54) |
| Month 8 of follow-up (Month 11 after primary infection/first-dose vaccination) | 11,644 | 10.89 (10.52 to 11.28) | 10,000 | 21.28 (20.79 to 21.78) | 0.52 (0.47 to 0.57) | 0.52 (0.47 to 0.57) |
| Natural infection versus mRNA-1273 vaccination study§ |  |  |  |  |  |  |
| Month 1 of follow-up (Month 4 after primary infection/first-dose vaccination) | 48,697 | 0.16 (0.13 to 0.19) | 48,694 | 0.20 (0.17 to 0.25) | 0.80 (0.60 to 1.07) | 0.80 (0.59 to 1.07) |
| Month 2 of follow-up (Month 5 after primary infection/first-dose vaccination) | 41,685 | 0.44 (0.38 to 0.50) | 41,580 | 0.79 (0.71 to 0.87) | 0.49 (0.40 to 0.61) | 0.48 (0.39 to 0.59) |
| Month 3 of follow-up (Month 6 after primary infection/first-dose vaccination) | 38,352 | 0.82 (0.74 to 0.91) | 38,149 | 1.50 (1.39 to 1.62) | 0.52 (0.43 to 0.63) | 0.50 (0.41 to 0.61) |
| Month 4 of follow-up (Month 7 after primary infection/first-dose vaccination) | 33,847 | 1.12 (1.02 to 1.22) | 33,615 | 2.29 (2.15 to 2.44) | 0.37 (0.29 to 0.46) | 0.37 (0.29 to 0.46) |
| Month 5 of follow-up (Month 8 after primary infection/first-dose vaccination) | 28,482 | 1.68 (1.55 to 1.81) | 28,028 | 3.86 (3.66 to 4.06) | 0.36 (0.30 to 0.42) | 0.35 (0.29 to 0.41) |
| Month 6 of follow-up (Month 9 after primary infection/first-dose vaccination) | 22,361 | 3.69 (3.48 to 3.91) | 21,512 | 8.00 (7.69 to 8.31) | 0.46 (0.41 to 0.51) | 0.44 (0.40 to 0.50) |
| Month 7 of follow-up (Month 10 after primary infection/first-dose vaccination) | 15,261 | 8.50 (8.14 to 8.87) | 14,016 | 15.91 (15.45 to 16.39) | 0.58 (0.53 to 0.62) | 0.57 (0.53 to 0.62) |
| Month 8 of follow-up (Month 11 after primary infection/first-dose vaccination) | 7,326 | 10.20 (9.79 to 10.62) | 6,436 | 18.35 (17.83 to 18.88) | 0.61 (0.52 to 0.72) | 0.61 (0.52 to 0.71) |

CI denotes confidence interval
\*A month was defined as 30 days.
<sup>†</sup>Cohorts were exact-matched in a 1:1 ratio by sex, 10-year age group, nationality, and timing of primary infection/first-dose vaccination to the first eligible individual with two-dose vaccination.
<sup>‡</sup>Cox regression analysis adjusted for sex, 10-year age group (Table 1), 10 nationality groups (Table 1), and timing of primary infection/first dose vaccination.
<sup>§</sup>No results were generated for months 1-3 after primary infection/first-dose vaccination. This is because a SARS-CoV-2 reinfection is conventionally defined as a documented infection $\geq 90$ days after an earlier infection, to avoid misclassification of prolonged PCR positivity as reinfections.<sup>6,8,9</sup> Therefore, cohorts were followed starting from $\geq 90$ (three months) after the documented primary infection/first-dose vaccination.

The overall adjusted hazard ratio for any severe, critical, or fatal COVID-19 was estimated at 0.32 (95% CI: 0.10-1.00; Table 2). The wide 95% CI reflected the rarity of COVID-19 hospitalizations in both the natural-infection and BNT162b2-vaccinated cohorts (Figure 1).

### Natural infection versus mRNA-1273 vaccination

The median time of follow-up was 143 days (IQR, 45-212 days) for the natural-infection cohort and 141 days (IQR, 45-206 days) for the mRNA-1273-vaccinated cohort (Figure 2B). A total of 2,376 reinfections were recorded in the natural-infection cohort during follow-up (Figure 1). Three of these reinfections progressed to severe COVID-19, but none to critical or fatal COVID-19. A total of 4,442 infections were recorded in the mRNA-1273-vaccinated cohort. Of these infections, 3 progressed to severe, 2 to critical, but none to fatal COVID-19.

Cumulative incidence of infection was estimated at 10.2% (95% CI: 9.8-10.6%) for the natural-infection cohort and at 18.4% (95% CI: 17.8-18.9%) for the mRNA-1273-vaccinated cohort, 240 days after the start of follow-up (Figure 2B). Incidence was limited in the natural-infection cohort until day 170 of follow-up, but then increased rapidly with the onset of the Omicron wave.^6,24,39,40^ Prior to day 170, incidence was dominated by the Alpha, Beta, and Delta variants.^3,20,39,41,42^ Meanwhile, there was considerable incidence in the mRNA-1273-vaccinated cohort prior to the Omicron wave, and much larger incidence after the onset of this wave.

The overall adjusted hazard ratio for infection was estimated at 0.51 (95% CI: 0.48-0.53; Table 2). However, the adjusted hazard ratio varied by month of follow-up (Table 3). It was 0.80 (95% CI: 0.59-1.07) in the first month of follow-up (fourth month after primary infection/vaccination), but decreased gradually to 0.35 (95% CI: 0.29-0.41) in the eighth month after primary infection/vaccination. Nonetheless, the adjusted hazard ratio increased after the onset of the Omicron wave. It was ∼0.60 during the time of follow-up that coincided with this wave.

The overall adjusted hazard ratio for any severe, critical, or fatal COVID-19 was estimated at 0.58 (95% CI: 0.14-2.43; Table 2). The wide 95% CI reflected the rarity of COVID-19 hospitalizations in both the natural-infection and mRNA-1273-vaccinated cohorts (Figure 1).

## Discussion

Natural infection was associated with an overall 50% lower incidence of SARS-CoV-2 infection than mRNA primary-series vaccination. Natural infection was also associated with lower incidence of COVID-19 hospitalizations than vaccination, but COVID-19 hospitalizations were rare for both the natural-infection and vaccinated cohorts. While vaccine protection against infection waned with time after the second dose, natural immunity demonstrated hardly any waning in protection for eight months of follow-up after the primary infection. However, onset of the Omicron wave led to a massive increase in incidence of reinfections in the natural-infection cohort and incidence of infections in the vaccinated cohorts. Yet, even during the Omicron wave, natural infection was still associated with 50% lower incidence than BNT162b2 vaccination, and 40% lower incidence than mRNA-1273 vaccination.

While natural infection was more protective than vaccine protection, it appears that initially, immediately after the second dose, the differences are minimal, if any. For the mRNA-1273 vaccine, the adjusted hazard ratio was 0.80 (95% CI: 0.59-1.07) in the fourth month after vaccination, indicating no statistically significant difference between protection of natural infection and that of mRNA-1273 vaccination. However, the divergence between the two protection types increased in subsequent months. Incidence among those with natural infection was 65% lower than that among those mRNA-1273-vaccinated in the eighth month after vaccination.

These findings may be explained by different roles for mucosal immunity in the protection.^14,15^ While natural infection was associated with stronger protection, vaccination remains the safest and optimal tool of protection against infection and COVID-19 hospitalization and death. Natural infection can lead to COVID-19 hospitalization and death at time of primary infection, and long COVID-19 after the infection, risks that are not present for vaccination. Protection of natural infection was compared to only primary-series vaccination. The differences in protection between natural infection and booster vaccination may be smaller.

The results of this study confirm findings that we reported recently. Protection of prior infection against reinfection with Omicron was estimated at 56.0%.^6^ Since BNT162b2 vaccine protection is negligible 6 or more months after the second dose,^43,44^ the adjusted hazard ratio of ∼0.50 during the Omicron wave implies ∼50% protection for natural infection against reinfection with Omicron. The study results confirmed the waning of protection of mRNA vaccines against pre-Omicron variants,^3,4^ and the lower vaccine protection against Omicron.^43,44^ The results also supported stronger protection and slower waning for mRNA-1273 protection than for BNT162b2 protection.^26,45^

This study has limitations. We investigated incidence of documented infections, but other infections may have occurred and gone undocumented, perhaps because of minimal/mild or no symptoms. While the vaccinated cohorts excluded those with a prior documented infection, some of those vaccinated may have experienced an undocumented prior infection. Therefore, the higher protection of natural infection compared to vaccination may have been underestimated. With the high and durable effectiveness of natural infection^5,6^ and mRNA primary-series vaccination^3,4^ against COVID-19 hospitalization and death and the young population of Qatar,^25^ case numbers were insufficient for precise estimation of differences in protection between natural infection and vaccination against COVID-19 hospitalization and death.

Depletion of the natural-infection cohort by COVID-19 mortality at time of primary infection may have biased this cohort toward healthier individuals with stronger immune responses. However, COVID-19 mortality has been low in Qatar’s predominantly young, working-age population,^25,46^ totaling 677 COVID-19 deaths (∼0.1% of primary infections) from the onset of the pandemic in early 2020 until March 14, 2022. Therefore, this seems unlikely to explain the study findings.

As an observational study, investigated cohorts were neither blinded nor randomized, so unmeasured or uncontrolled confounding cannot be excluded. While matching was done for sex, age, nationality, and timing of prior infection/first dose, this was not possible for other factors, such as comorbidities, occupation, or geography, as such data were unavailable. However, matching was done to control for factors that affect infection exposure in Qatar.^25,27-30^ Matching by age may have reduced potential bias due to comorbidities. The number of individuals with severe chronic conditions is also small in Qatar’s young population.^25,46^ Matching by nationality may have partially controlled for differences in occupational risk or socio-economic status, given the statistical association between the nationality of workers and occupation type in Qatar.^25,27-30^ Qatar is essentially a city state and infection incidence and vaccination were broadly distributed across neighborhoods/areas. That is, geography is not likely to have been a confounding factor. Lastly, matching by the considered factors has been shown to provide adequate control of bias in prior studies that used control groups in Qatar.^3,4,19,26,31^ These included unvaccinated cohorts versus vaccinated cohorts within two weeks of the first dose,^3,4,19,31^ when vaccine protection is negligible,^1,2^ and mRNA-1273-versus BNT162b2-vaccinated cohorts, also in the first two weeks after the first dose.^26^

In conclusion, natural infection was associated with lower incidence of SARS-CoV-2 infection and COVID-19 hospitalizations, regardless of the variant, than mRNA primary-series vaccination. While differences between natural-infection protection and vaccine protection were minimal immediately after the second vaccine dose, divergence between the two types of protection occurred in subsequent months, consistent with waning of vaccine immunity, but very slow waning in natural-infection immunity, at least against pre-Omicron variants. While natural infection was associated with stronger protection, vaccination remains the safest and optimal tool of protection against infection and COVID-19 hospitalization and death.

## Data Availability

The dataset of this study is a property of the Qatar Ministry of Public Health that was provided to the researchers through a restricted-access agreement that prevents sharing the dataset with a third party or publicly. Future access to this dataset can be considered through a direct application for data access to Her Excellency the Minister of Public Health (https://www.moph.gov.qa/english/Pages/default.aspx). Aggregate data are available within the manuscript and its Supplementary information.

## Sources of support and acknowledgements

We acknowledge the many dedicated individuals at Hamad Medical Corporation, the Ministry of Public Health, the Primary Health Care Corporation, Qatar Biobank, Sidra Medicine, and Weill Cornell Medicine-Qatar for their diligent efforts and contributions to make this study possible.

The authors are grateful for institutional salary support from the Biomedical Research Program and the Biostatistics, Epidemiology, and Biomathematics Research Core, both at Weill Cornell Medicine-Qatar, as well as for institutional salary support provided by the Ministry of Public Health, Hamad Medical Corporation, and Sidra Medicine. The authors are also grateful for the Qatar Genome Programme and Qatar University Biomedical Research Center for institutional support for the reagents needed for the viral genome sequencing. The funders of the study had no role in study design, data collection, data analysis, data interpretation, or writing of the article. Statements made herein are solely the responsibility of the authors.

## Author contributions

HC co-designed the study, performed the statistical analyses, and co-wrote the first draft of the article. LJA conceived and co-designed the study, led the statistical analyses, and co-wrote the first draft of the article. HY, HAK, and MS conducted viral genome sequencing. PT and MRH conducted the multiplex, RT-qPCR variant screening and viral genome sequencing. All authors contributed to data collection and acquisition, database development, discussion and interpretation of the results, and to the writing of the manuscript. All authors have read and approved the final manuscript.

## Supplementary Appendix

### Section S1. Laboratory methods and variant ascertainment

#### Real-time reverse-transcription polymerase chain reaction testing

Nasopharyngeal and/or oropharyngeal swabs were collected for polymerase chain reaction (PCR) testing and placed in Universal Transport Medium (UTM). Aliquots of UTM were: 1) extracted on KingFisher Flex (Thermo Fisher Scientific, USA), MGISP-960 (MGI, China), or ExiPrep 96 Lite (Bioneer, South Korea) followed by testing with real-time reverse-transcription PCR (RT-qPCR) using TaqPath COVID-19 Combo Kits (Thermo Fisher Scientific, USA) on an ABI 7500 FAST (Thermo Fisher Scientific, USA); 2) tested directly on the Cepheid GeneXpert system using the Xpert Xpress SARS-CoV-2 (Cepheid, USA); or 3) loaded directly into a Roche cobas 6800 system and assayed with the cobas SARS-CoV-2 Test (Roche, Switzerland). The first assay targets the viral S, N, and ORF1ab gene regions. The second targets the viral N and E-gene regions, and the third targets the ORF1ab and E-gene regions.

All PCR testing was conducted at the Hamad Medical Corporation Central Laboratory or Sidra Medicine Laboratory, following standardized protocols.

#### Rapid antigen testing

Severe acute respiratory syndrome coronavirus 2 (SARS-CoV-2) antigen tests were performed on nasopharyngeal swabs using one of the following lateral flow antigen tests: Panbio COVID-19 Ag Rapid Test Device (Abbott, USA); SARS-CoV-2 Rapid Antigen Test (Roche, Switzerland); Standard Q COVID-19 Antigen Test (SD Biosensor, Korea); or CareStart COVID-19 Antigen Test (Access Bio, USA). All antigen tests were performed point-of-care according to each manufacturer’s instructions at public or private hospitals and clinics throughout Qatar with prior authorization and training by the Ministry of Public Health (MOPH). Antigen test results were electronically reported to the MOPH in real time using the Antigen Test Management System which is integrated with the national Coronavirus Disease 2019 (COVID-19) database.

#### Classification of infections by variant type

Surveillance for SARS-CoV-2 variants in Qatar is based on viral genome sequencing and multiplex RT-qPCR variant screening^1^ of random positive clinical samples,^2-7^ complemented by deep sequencing of wastewater samples.^4,8^ Further details on the viral genome sequencing and multiplex RT-qPCR variant screening throughout the SARS-CoV-2 waves in Qatar can be found in previous publications.^2-7,9-12^

### Section S2. COVID-19 severity, criticality, and fatality classification

Severe COVID-19 disease was defined per the World health Organization (WHO) classification as a SARS-CoV-2 infected person with “oxygen saturation of <90% on room air, and/or respiratory rate of >30 breaths/minute in adults and children >5 years old (or ≥60 breaths/minute in children <2 months old or ≥50 breaths/minute in children 2-11 months old or ≥40 breaths/minute in children 1–5 years old), and/or signs of severe respiratory distress (accessory muscle use and inability to complete full sentences, and, in children, very severe chest wall indrawing, grunting, central cyanosis, or presence of any other general danger signs)”.^13^ Detailed WHO criteria for classifying SARS-CoV-2 infection severity can be found in the WHO technical report.^13^

Critical COVID-19 disease was defined per WHO classification as a SARS-CoV-2 infected person with “acute respiratory distress syndrome, sepsis, septic shock, or other conditions that would normally require the provision of life sustaining therapies such as mechanical ventilation (invasive or non-invasive) or vasopressor therapy”.^13^ Detailed WHO criteria for classifying SARS-CoV-2 infection criticality can be found in the WHO technical report.^13^

COVID-19 death was defined per WHO classification as “a death resulting from a clinically compatible illness, in a probable or confirmed COVID-19 case, unless there is a clear alternative cause of death that cannot be related to COVID-19 disease (e.g. trauma). There should be no period of complete recovery from COVID-19 between illness and death. A death due to COVID-19 may not be attributed to another disease (e.g. cancer) and should be counted independently of preexisting conditions that are suspected of triggering a severe course of COVID-19”. Detailed WHO criteria for classifying COVID-19 death can be found in the WHO technical report.^14^

**Table S1.** STROBE checklist for cohort studies.

|  | Item No | Recommendation | Main Text page |
| --- | --- | --- | --- |
| **Title and abstract** | 1 | (a) Indicate the study’s design with a commonly used term in the title or the abstract<br>(b) Provide in the abstract an informative and balanced summary of what was done and what was found | Abstract<br>Abstract |
| **Introduction** |  |  |  |
| Background/rationale | 2 | Explain the scientific background and rationale for the investigation being reported | Introduction |
| Objectives | 3 | State specific objectives, including any prespecified hypotheses | Introduction |
| **Methods** |  |  |  |
| Study design | 4 | Present key elements of study design early in the paper | Methods (‘Study design’) |
| Setting | 5 | Describe the setting, locations, and relevant dates, including periods of recruitment, exposure, follow-up, and data collection | Methods (‘Study design’) & Figure 1 |
| Participants | 6 | (a) Give the eligibility criteria, and the sources and methods of selection of participants. Describe methods of follow-up<br>(b) For matched studies, give matching criteria and number of exposed and unexposed | Methods (‘Study design’) & Figure 1 |
| Variables | 7 | Clearly define all outcomes, exposures, predictors, potential confounders, and effect modifiers. Give diagnostic criteria, if applicable | Methods (‘Study design’ & ‘Study outcomes’), Table 1, & Sections S1& S2 in Supplementary Appendix |
| Data sources/measurement | 8* | For each variable of interest, give sources of data and details of methods of assessment (measurement). Describe comparability of assessment methods if there is more than one group | Methods (‘Study population and data sources’ & ‘Statistical analysis’, paragraph 1), Table 1, & Sections S1 & S2 in Supplementary Appendix |
| Bias | 9 | Describe any efforts to address potential sources of bias | Methods (‘Study design’, paragraphs 2-4) |
| Study size | 10 | Explain how the study size was arrived at | Figure 1 |
| Quantitative variables | 11 | Explain how quantitative variables were handled in the analyses. If applicable, describe which groupings were chosen and why | Methods (‘Study design’, paragraphs 3 and 5) & Table 1 |
| Statistical methods | 12 | (a) Describe all statistical methods, including those used to control for confounding | Methods (‘Statistical analysis’) |
|  |  | (b) Describe any methods used to examine subgroups and interactions | Methods (‘Statistical analysis’, paragraph 2) |
|  |  | (c) Explain how missing data were addressed | NA, see Methods (‘Study population and data sources’, paragraph 1) |
|  |  | (d) If applicable, explain how loss to follow-up was addressed | NA, see Methods (‘Study design’, paragraph 1) |
|  |  | (e) Describe any sensitivity analyses | NA |
| **Results** |  |  |  |
| Participants | 13* | (a) Report numbers of individuals at each stage of study—eg numbers potentially eligible, examined for eligibility, confirmed eligible, included in the study, completing follow-up, and analysed<br>(b) Give reasons for non-participation at each stage<br>(c) Consider use of a flow diagram | Results (‘Study population for BNT162b2 vaccine’ & ‘Study population for mRNA-1273 vaccine), Figure 1, & Table 1 |
| Descriptive data | 14 | (a) Give characteristics of study participants (eg demographic, clinical, social) and information on exposures and potential confounders | Results (‘Study population’), Table 1, & Figures S1 & S2 in Supplementary Appendix |
|  |  | (b) Indicate number of participants with missing data for each variable of interest | NA, see Methods (‘Study population and data sources’, paragraph 1) |
|  |  | (c) Summarise follow-up time (eg, average and total amount) | Results (‘Natural infection versus BNT162b2 vaccination’, paragraph 1 & ‘Natural infection versus mRNA-1273 vaccination’, paragraph 1), Figure 2, & Table 2 |
| Outcome data | 15 | Report numbers of outcome events or summary measures over time | Results (‘Natural infection versus BNT162b2 vaccination’ & ‘Natural infection versus mRNA-1273 vaccination’), Figures 1-2, & Table 2 |
| Main results | 16 | (a) Give unadjusted estimates and, if applicable, confounder-adjusted estimates and their precision (eg, 95% confidence interval). Make clear which confounders were adjusted for and why they were included | Results ('Effectiveness of BNT162b2 booster against Results ('Natural infection versus BNT162b2 vaccination' & 'Natural infection versus mRNA-1273 vaccination'), Figure 2, & Table 2 |
|  |  | (b) Report category boundaries when continuous variables were categorized | Table 1 |
|  |  | (c) If relevant, consider translating estimates of relative risk into absolute risk for a meaningful time period | NA |
| Other analyses | 17 | Report other analyses done—eg analyses of subgroups and interactions, and sensitivity analyses | Results ('Effectiveness of BNT162b2 booster against Results ('Natural infection versus BNT162b2 vaccination', paragraph 3 & 'Natural infection versus mRNA-1273 vaccination', paragraph 3), & Table 3 |
| Discussion |  |  |  |
| Key results | 18 | Summarise key results with reference to study objectives | Discussion, paragraphs 1-3 |
| Limitations | 19 | Discuss limitations of the study, taking into account sources of potential bias or imprecision. Discuss both direction and magnitude of any potential bias | Discussion, paragraphs 4-5 |
| Interpretation | 20 | Give a cautious overall interpretation of results considering objectives, limitations, multiplicity of analyses, results from similar studies, and other relevant evidence | Discussion, paragraph 6 |
| Generalisability | 21 | Discuss the generalisability (external validity) of the study results | Discussion, paragraphs 4-5 and Table S2 in Supplementary Appendix |
| Other information |  |  |  |
| Funding | 22 | Give the source of funding and the role of the funders for the present study and, if applicable, for the original study on which the present article is based | Sources of support & acknowledgements |

**Figure S1.**
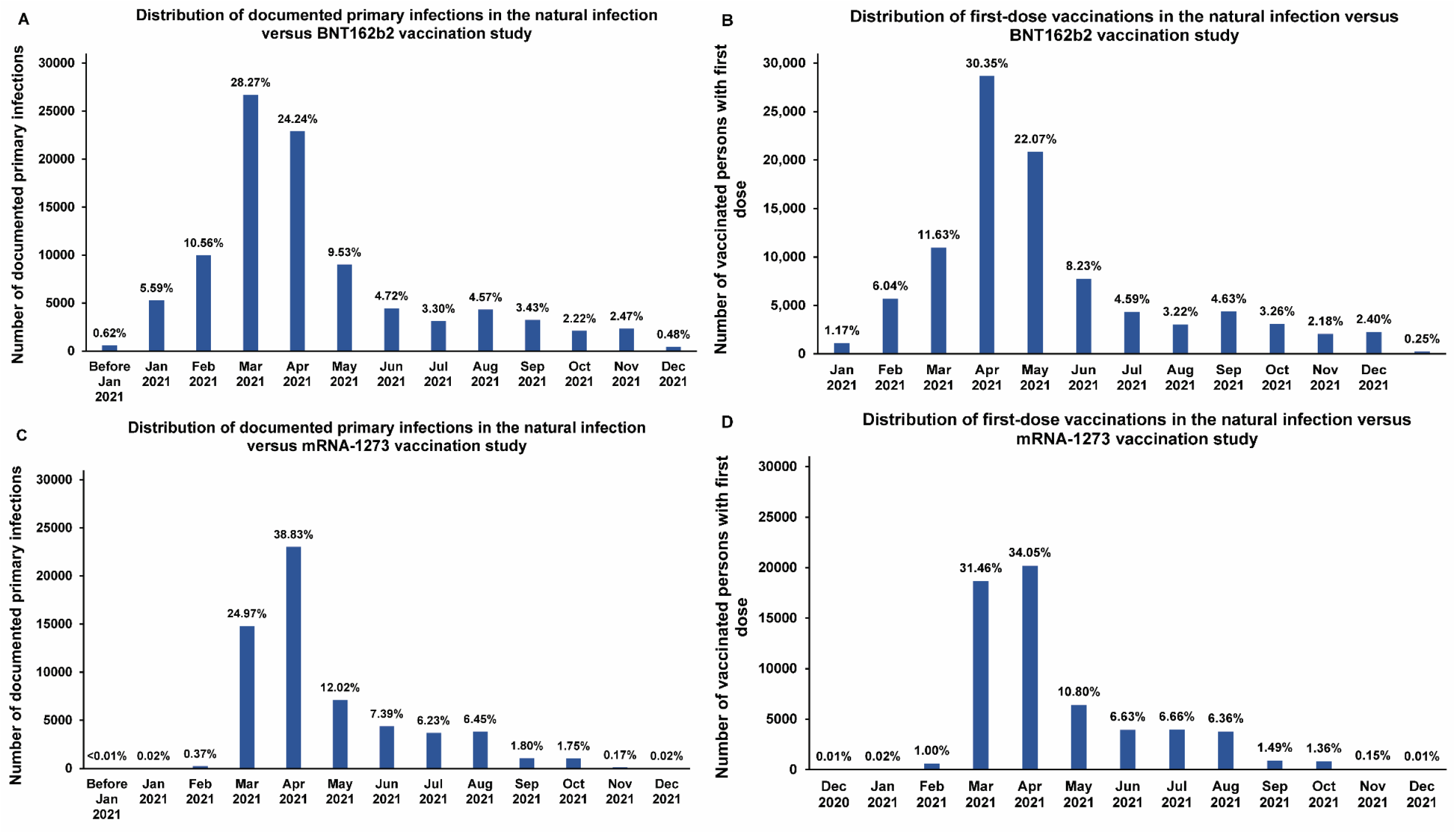
Distribution of documented SARS-CoV-2 primary infections and of first-dose vaccinations by calendar month in the matched cohorts of the natural-infection-versus-BNT162b2-vaccination study (panels A and B) and the natural-infection-versus-mRNA-1273-vaccination study (panels C and D).

**Figure S2.**
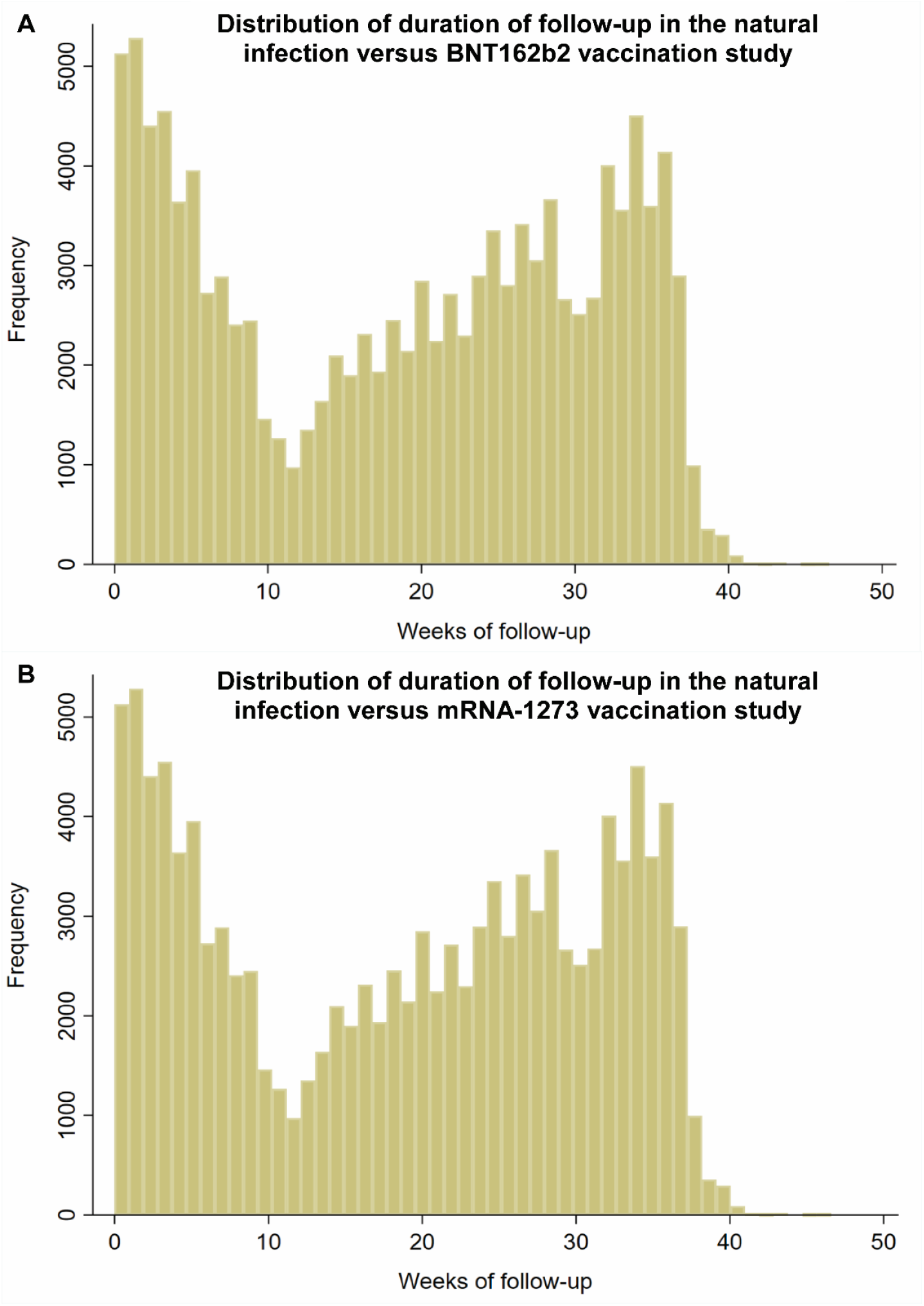
Distribution of the durations of follow-up in the matched cohorts of the A) natural-infection-versus-BNT162b2-vaccination study and B) natural-infection-versus-mRNA-1273-vaccination study.

**Table S2.** Representativeness of study participants.

|  |  |
| --- | --- |
| **Category** |  |
| Disease, problem, or condition under investigation | Protection conferred by natural SARS-CoV-2 infection versus BNT162b2 and mRNA-1273 vaccination against SARS-CoV-2 infection and against COVID-19 hospitalization and death. |
| Special considerations related to |  |
| Sex and gender | Two national matched, retrospective target-trial cohort studies were conducted to compare incidence of SARS-CoV-2 infection and COVID-19 hospitalization and death among those with a documented primary infection to incidence among those with a two-dose primary-series mRNA vaccination. Cohorts were exact-matched by sex to control for potential differences in the risk of exposure to SARS-CoV-2 infection by sex. |
| Age | Cohorts were exact-matched by 10-year age group to control for potential differences in the risk of exposure to SARS-CoV-2 infection by age. Nonetheless, with the young population of Qatar, our findings may not be generalizable to other countries where elderly citizens constitute a larger proportion of the total population. |
| Race or ethnicity group | Cohorts were exact-matched by nationality to control for potential differences in the risk of exposure to SARS-CoV-2 infection by nationality. Nationality is associated with race and ethnicity in the population of Qatar. |
| Geography | Individual-level data on geography were not available, but Qatar is essentially a city state and infection incidence and vaccination was broadly distributed across the country's neighborhoods/areas. Cohorts were exact-matched by nationality to control for potential differences in the risk of exposure to SARS-CoV-2 infection by nationality. Qatar has unusually diverse demographics in that 89% of the population are international expatriate residents coming from over 150 countries from all world regions. |
| Other considerations | Individual-level data on co-morbid conditions were not available, but only a small proportion of the study population may have had serious co-morbid conditions. Only 9% of the population of Qatar are $\geq 50$ years of age (older age as proxy for co-morbidities). The national list of persons prioritized to receive the vaccine during the first phase of vaccine roll-out included only 19,800 individuals of all age groups with serious co-morbid conditions. Individual-level data on occupation were not available but matching by nationality may have (partially) controlled the differences in occupational risk, in consideration of the association between nationality and occupation in Qatar. To control for time since inducement of SARS-CoV-2 immunity, each individual in the vaccinated cohorts was matched to an individual in the natural-infection cohort who had her/his documented infection within a week after the vaccinated match received the first vaccine dose. |
| Overall representativeness of this study | The study was based on the total population of Qatar and thus the study population is broadly representative of the diverse, by national background, but young and predominantly male, total population of Qatar. While there could be differences in the risk of exposure to SARS-CoV-2 infection by sex, age, and nationality, cohorts were exact-matched by these factors to control for their potential impact on our estimates. Given that only 9% of the population of Qatar are $\geq 50$ years of age and the limited proportion of the population with significant co-morbidities, our estimates may not be generalizable to other countries where elderly citizens constitute a larger proportion of the total population or where co-morbid conditions are prevalent. |

COVID-19 coronavirus disease 2019, and SARS-CoV-2 severe acute respiratory syndrome coronavirus 2.

